# Low-to-moderate dose ionizing radiation and cerebrovascular disease mortality in U.S. radiologic technologists, 1983-2021

**DOI:** 10.64898/2026.10.21.26366178

**Authors:** Mark P. Little, Martha S. Linet, Isabelle F. van der Velpen, Taeeun Kwon, Hae Gin Han, Choonsik Lee, Elizabeth K. Cahoon, Jim Z. Mai, Jeremy S. Miller, Bruce H. Alexander, Cari M. Kitahara

**Affiliations:** Radiation Epidemiology Branch, Division of Cancer Epidemiology and Genetics, National Cancer Institute, NIH, DHHS, Rockville, MD 20892-9778, MD, USA; Information Management Services, Silver Spring, MD 20904, USA; Division of Environmental Health Sciences, School of Public Health, University of Minnesota, Minneapolis, MN 55409, USA

**Keywords:** ionizing radiation, cerebrovascular disease, ischemic stroke, hemorrhagic stroke, cardiovascular disease risk factors, cohort study, medical workers, occupational epidemiology

## Abstract

**Background:** Substantial data confirm the association of cerebrovascular disease (CeVD) with high-dose (>1 Gy) ionizing radiation exposures, but less is known about risks at low- to moderate doses (<1 Gy). A few studies have reported increased CeVD risks at doses <500 mGy, but none of these provided data on CeVD risks by subtype. Few studies have investigated the potential modifying effects of age, sex, and other major CeVD risk factors.

**Methods:** 110,368 radiologic technologists certified for ≥2 years during 1926-1982 with at least one baseline survey response (1983-1998) were followed-up through December 2021. Linear proportional hazards regression analysis was performed of cumulative occupational radiation exposure and CeVD mortality risks, using estimated thyroid-absorbed dose as a proxy for carotid artery dose, in addition to brain and badge dose.

**Results:** During a median 35.1 (maximum 38.9) years of follow-up, 1,199 deaths from CeVD (634 ischemic stroke, 369 hemorrhagic stroke, 196 other types) were identified. Cumulative thyroid dose (mean/maximum=68.4/2784 mGy) was positively associated with deaths from overall CeVD (excess hazard ratio [EHR]/100 mGy=0.07 95% CI 0.01, 0.16, *p*=0.01) and ischemic stroke (EHR/100 mGy=0.10, 95% CI 0.02, 0.23, *p*=0.01), while no clear associations were observed for hemorrhagic stroke (EHR/100 mGy=0.04, 95% CI −0.04, 0.23 *p*=0.43) or other CeVD (EHR/100 mGy=0.02, 95% CI −0.10, 0.29, *p*=0.81). Trends were similar for total CeVD using badge dose (EHR/100 mSv=0.08, 95% CI 0.02, 0.16, *p*<0.01), but larger with brain dose (EHR/100 mGy=0.34, 95% CI −0.00, 0.81, *p*=0.05). Body mass index modified the CeVD dose-response (*p*=0.04), but year of birth, time since exposure, age at exposure, attained age, sex, smoking, and alcohol consumption did not modify the CeVD dose-response (all *p*>0.05), nor did adjusting for history of diabetes, hypertension or high cholesterol. Trends remained significant when restricting thyroid doses to <1,000 mGy (EHR/100 mGy=0.06, 95% CI −0.00, 0.16, 1-sided *p*=0.03).

**Conclusions:** In the first cohort study to assess low-to-moderate occupational ionizing radiation dose in medical workers with CeVD mortality risk by subtype, greater occupational ionizing radiation exposure was associated with increased mortality risk from total CeVD and ischemic stroke, independent of demographic and lifestyle-related factors and comorbid conditions.

**Clinical Perspective:**

- For the first time we have documented radiation-related excess cerebrovascular disease risk, specifically of ischemic type, in a group of US radiologic technologists at relatively modest levels of dose (<1000 mGy).
- Unique among occupational cohorts there is rich data on lifestyle and other risk factors (including smoking, alcohol consumption, body mass index, diabetes, elevated serum cholesterol, or hypertension) in this group, few of which materially modify the radiation dose response.
- These results reinforce the need for radiation protection practices aimed at keeping exposures to medical staff and patients as low as reasonably achievable.

## Introduction

Cerebrovascular disease (CeVD) is the fourth leading cause of death in the USA, accounting for approximately 160,000 deaths annually (about 5.4% of all deaths).^1^ Ischemic stroke comprises the majority (87%) of the approximately 800,000 new CeVD diagnoses each year. Established risk factors include older age, excess body weight, smoking, alcohol consumption, diabetes, hyperlipidaemia, and hypertension.^2^

Substantial data confirm the association of CeVD with high-dose (>1 Gy) ionizing radiation exposures, ^3^ but less is known about risks in the low- to moderate-dose range (<1 Gy). The International Commission on Radiological Protection, based on older epidemiological and experimental data, classifies CeVD (and other cardiovascular diseases) as a tissue reaction effect with a threshold dose of 0.5 Gy below which no effects are anticipated.^4^ Despite inconsistent findings across all studies in this range,^3^ a few recent cohorts have reported increased CeVD risks at levels below 0.5 Gy,^5,6^ but none of these provided data on CeVD risks by subtype. Few studies have investigated the potential modifying effects of age, sex, and other major CeVD risk factors.

Therefore, we used data from the large nationwide U.S. Radiologic Technologists (USRT) cohort study to investigate the association of low- to moderate-dose, protracted radiation exposure with overall CeVD mortality and major CeVD subtypes (ischemic, hemorrhagic, all other). In addition to nearly 40 years of mortality follow-up, the cohort includes comprehensive information on demographic, lifestyle, and medical history factors and a state-of-the-art occupational radiation dose reconstruction spanning over 100 years of work history.^7^ In addition to evaluating the magnitude and shape of the dose-response, we examined potential confounding and modifying effects of age, sex, and other CeVD risk factors, including obesity, smoking, alcohol consumption, and medical history of diabetes, high cholesterol, and hypertension.

## Methods

### Study population and follow-up

#### Overview

The USRT study population and methods are described elsewhere (see also www.radtechstudy.nci.nih.gov).^7–9^ Briefly, the U.S. National Cancer Institute, in collaboration with the University of Minnesota and the American Registry of Radiologic Technologists (ARRT), is studying cancer and other serious disease incidence and mortality among 146,022 (106,953 female) U.S. radiologic technologists who were certified for at least two years during 1926-1982. Annual follow-up was conducted through yearly re-certification with the ARRT. For those who did not recertify, vital status and the cause of death through December 31, 2021 were determined through linkage with the Social Security Administration (up to 2013) and National Death Index (NDI-*Plus*) records.

#### Data collection

Four cohort follow-up surveys were conducted using mailed questionnaires. The first questionnaire (1983-89) was mailed to 132,298 radiologic technologists identified as alive and with a valid address, of whom 90,305 (68%) responded. The second questionnaire (1994-98) was mailed to 126,628 living and located technologists, of whom 90,972 (72%) responded. The third questionnaire (2003-2005) was mailed or administered by telephone to 101,694 living cohort members (administration was by telephone to technologists first working before 1950) who had completed at least one of the two prior surveys; 73,838 technologists (73%) responded. The fourth questionnaire was sent to 93,787 living and located cohort members in 2012-2013, with 62% response rate.

#### Study population and follow-up

Technologists eligible for the mortality analysis had to have completed the first or the second survey questionnaire and were followed from their first completed questionnaire through the earlier of date of death, age 90, or December 31, 2021. A cutoff of age 90 was used to address the possible unreliability of cause of death ascertainment at advanced ages, also possible loss of follow-up via emigration (although this was not expected to be substantial).^10^ Of the 110,373 technologists meeting these criteria, the date of last vital status was unknown for five, leaving an analysis dataset of 110,368 technologists.

### Disease ascertainment and classification

As above, date and underlying cause of death was derived from NDI-*Plus*. We use deaths coded as either ICD9 430-438 or ICD10 I60-69 for overall CeVD mortality. The ICD9 and ICD10 codes for the main types of CeVD mortality (ischemic disease, hemorrhagic disease, all other types of CeVD) are as given in Table 3.

### Dose estimation

Individual doses were reconstructed using the recently developed U.S. Radiologic Technologists Dosimetry System 2025 (URDS25), which estimates historical badge doses and subsequently converts them to organ-specific absorbed doses. The badge dose estimation methods are described in detail in Kwon *et al.*^7^ Briefly, badge doses were estimated for USRT technologists who completed one or more self-administered questionnaires. Because complete badge dose measurements covered only 43% of the occupational radiation exposure of the USRT cohort members and badge measurements were limited before the late 1970s, the badge dose reconstruction used: (1) individual badge measurements (mostly covering 1978–2020), (2) detailed questionnaire-based work history and radiation protection measures and (3) historical literature on radiologic exposures to medical radiation workers (covering 1916– 1955). We developed probability density functions of badge dose by decade from which each missing dose was sampled. The estimated annual technologist population mean badge doses decreased substantially from over 100 mSv up to the mid-1930s, to under 1 mSv in the 2000s.^7^

Thyroid dose was used as a surrogate for carotid dose, to reflect the pathophysiological relevance of carotid atherosclerotic disease in CeVD risk,^11,12^ and following a previous approach used to model dose-response for circulatory disease.^13^ Full brain doses were also used to assess exposure directly to the brain tissue and vasculature. To estimate absorbed doses to the thyroid and brain, the badge dose estimate for each person-year was first apportioned among diagnostic radiography, routine fluoroscopy, nuclear medicine, and fluoroscopically-guided interventional procedures using questionnaire-derived procedure and work-history information. These procedure-specific contributions were then converted to thyroid and brain doses using conversion coefficients that accounted for relevant exposure characteristics, including the radiation energy spectrum and exposure geometry.^14,15^ The conversion coefficients were calculated using a comprehensive library of 193 adult female and male computational anatomical phantoms covering a wide range of body sizes and were individualized by interpolation based on each technologist’s height and weight.^16,17^ For fluoroscopically guided interventional procedures, questionnaire-derived probabilities of thyroid-shield use were used to weight thyroid dose coefficients with and without shield attenuation. All stages, from badge dose reconstruction through organ dose conversion, were performed probabilistically using 1,000 Monte Carlo realizations to account for multiple sources of uncertainty, including those associated with badge dose estimates, procedure-specific dose contributions, thyroid-shield use probabilities, and systematic variation in dose conversion coefficients. The doses reflect technologists’ exposures from external diagnostic and therapeutic radiological procedures, and were mostly from X-rays.^7^

### Cardiovascular risk factor information

The following questionnaire-derived variables were selected *a priori* as adjusting covariates in most regression models because of their known associations with CeVD,^18,19^ and potential for confounding or mediating the relationship between radiation exposure and CeVD mortality. These variables included sex, race, Hispanic ethnicity, birth year, diabetes, hypertension, elevated serum cholesterol, BMI, baseline smoking status (current smoker/ex-smoker/never smoker, pack years of cigarette smoking), and baseline alcohol status (number of alcoholic drinks per week). Data for the *a priori*-specified potential confounding variables were obtained from the first of the two baseline questionnaires, while the data on potential mediators (diabetes, hypertension, elevated cholesterol) were reported for the first time in the second questionnaire.

### Statistical Methods

Associations of occupational radiation exposure and CeVD mortality were assessed using Cox proportional hazards models,^20^ with age as timescale. We evaluated several dose metrics including thyroid dose, whole brain dose, and estimated badge dose, each lagged by 10 years, as has previously been done in analysis of this endpoint.^5,21^ We assessed potential modifications to the dose trend by time since exposure, age at exposure and attained age, all of these defined using moving time windows with dose accumulated accordingly. We explored adding adjustment for, and modification to the dose-response for certain medical and lifestyle variables listed above. Diabetes, elevated serum cholesterol and hypertension may be on the pathway between ionizing radiation exposure and CeVD mortality and may also, by virtue of their appearance on second, but not first, mailed questionnaires, be markers of survival to the later periods of follow-up; analyses using these were therefore restricted to those answering the second questionnaire (LQ2) and starting follow-up at the date of completion of the questionnaire. Missing values in factor variables (all the variables used for adjustment, which included BMI) were dealt with by adding levels to code for missingness. Further details on statistical methods, including further sensitivity analysis, are given in Supplement B. Analysis was conducted using R ^22^ and Fortran 95-2003 code written by the first author.

## Results

The study population comprises 110,368 persons, 83,717 (75.9%) of whom were female (Table 1). Mean follow-up was 31.2 years per person (median = 35.1, maximum = 38.9), amounting to a total of 3,447,039 person years of follow-up. The mean age at entry to follow-up was 41.1 years, and the mean age at exit 72.3 years. Of the cohort of 110,368 persons, 27,845 had died by the end of follow-up, of which 1376 deaths (4.9%) were due to CeVD; for follow-up restricted to age 90 there were 25,531 deaths, and of these 1199 (4.7%) occurred before the age of 90. The mean cumulative thyroid dose (at the end of follow-up) was 68.4 mGy, the mean film badge dose 78.8 mSv, and the mean brain dose 14.3 mGy. The cohort was predominantly White (94.2%) and of non-Hispanic ethnicity (97.1%).

**Table 1.** Descriptive data for USRT cerebrovascular disease (CeVD) data. Data restricted to those with known end of follow-up, and date of end of follow-up exceeding date of start of follow-up.

|  | Mean | Median | Minimum | Maximum |
| --- | --- | --- | --- | --- |
| Birth year | 1946.4 | 1949.0 | 1890.7 | 1966.5 |
| Age at entry | 41.1 | 38.6 | 22.3 | 90.0 |
| Age at exit | 72.3 | 71.3 | 25.8 | 90.0 |
| Year at entry | 1987.5 | 1985.4 | 1980.7 | 1998.9 |
| Year at exit | 2018.7 | 2022.0 | 1980.7 | 2022.0 |
| Years of follow-up | 31.2 | 35.1 | 0.0 | 38.9 |
| Mean thyroid dose, mGy<sup>a</sup> | 68.42 | 42.02 | 0.00 | 2784.19 |
| Mean brain dose, mGy<sup>a</sup> | 14.27 | 9.06 | 0.00 | 409.46 |
| Mean film badge dose, mSv<sup>a</sup> | 78.77 | 48.11 | 0.00 | 2990.10 |
| Total number of deaths / deaths restricted to age ≤ 90 | 27,845 / 25,531 |  |  |  |
| Number of CeVD deaths / deaths restricted to age ≤ 90 | 1376 / 1199 |  |  |  |
| Hemorrhagic CeVD deaths / deaths restricted to age ≤ 90 | 387 / 369 |  |  |  |
| Ischemic CeVD deaths / deaths restricted to age ≤ 90 | 745 / 634 |  |  |  |
| Other CeVD deaths / deaths restricted to age ≤ 90 | 244 / 196 |  |  |  |
| Number of person years follow-up | 3,447,039 |  |  |  |
| Number of persons | 110,368 |  |  |  |
|  | Number | Percentage |  |  |
| Males | 26,651 | 24.1 |  |  |
| Females | 83,717 | 75.9 |  |  |
| White | 103,922 | 94.2 |  |  |
| Black | 3544 | 3.2 |  |  |
| Asian/PI | 1278 | 1.2 |  |  |
| American Indian/Alaska Natives | 302 | 0.3 |  |  |
| Other | 1204 | 1.1 |  |  |
| Unknown | 118 | 0.1 |  |  |
| Non-Hispanic ethnicity | 107,203 | 97.1 |  |  |
| Hispanic ethnicity | 2707 | 2.5 |  |  |
| Ethnicity unknown | 458 | 0.4 |  |  |

<sup>a</sup>unlagged dose at end of follow-up

Table 2 shows the univariable associations for various baseline variables with CeVD mortality, most of which were highly significant (*p*<0.001), except for race, ethnicity, and elevated serum cholesterol (the latter restricted to LQ2 follow-up). Risks were higher for males, earlier years of birth, current smokers, persons smoking >15 pack-years, persons consuming ≥15 alcoholic drinks per week, persons with a BMI ≥30 kg m^-2^, persons with diabetes and persons with hypertension (Table 2). Supplement A Table A1 and Supplement B details the results of a stepwise multivariable analysis, variables being added in order of the significance in the univariable analysis of Table 2, demonstrating the highly significant effect on baseline risk of baseline smoking status, baseline smoking pack years, birth year, baseline BMI and sex (all *p*<0.005), and to a lesser extent of baseline alcohol consumption (*p*=0.043). All these covariates were used as stratifying variables in most dose-response analyses.

**Table 2.** Univariable analysis of baseline variables in relation to cerebrovascular disease (CeVD) using a Cox proportional hazards model with age as timescale.

| Variable | Persons | CeVD deaths | Hazard ratio (95% CI) | Heterogeneity *p*-value |
| --- | --- | --- | --- | --- |
| *All CeVD* | 110,368 | 1199 |  |  |
| *Sex* |  |  |  |  |
| Female | 83,717 | 823 | 1 = reference | 5.80 x 10<sup>-6</sup> |
| Male | 26,651 | 376 | 1.33 (1.18, 1.51) |  |
| *Birth year* |  |  |  |  |
| <1909 | 623 | 32 | 3.37 (2.17, 5.25) | 4.91 x 10<sup>-12</sup> |
| 1910-1919 | 2523 | 139 | 3.17 (2.32, 4.33) |  |
| 1920-1929 | 6061 | 245 | 2.27 (1.71, 3.02) |  |
| 1930-1939 | 14,936 | 378 | 1.78 (1.37, 2.32) |  |
| 1940-1949 | 36,256 | 293 | 1.48 (1.17, 1.89) |  |
| 1950-1959 | 49,108 | 111 | 1 = reference |  |
| 1960+ | 861 | 1 | 0.86 (0.12, 6.18) |  |
| *Race* |  |  |  |  |
| White | 103,922 | 1111 | 1 = reference | 0.533 |
| Black | 3544 | 54 | 1.22 (0.93, 1.61) |  |
| Asian/Pacific Islander | 1278 | 14 | 0.84 (0.49, 1.42) |  |
| American Indian/Alaska native | 302 | 3 | 1.02 (0.33, 3.17) |  |
| Other | 1204 | 16 | 1.29 (0.79, 2.12) |  |
| Unknown | 118 | 1 | 0.48 (0.07, 3.38) |  |
| *Hispanic ethnicity* |  |  |  |  |
| Non-Hispanic | 107,203 | 1163 | 1 = reference | 0.732 |
| Hispanic | 2707 | 31 | 1.07 (0.75, 1.52) |  |
| Unknown | 458 | 5 | 0.74 (0.31, 1.78) |  |
| *Baseline smoking status* |  |  |  |  |
| Never smoked | 52,265 | 431 | 1 = reference | <2.22 x 10<sup>-16</sup> |
| Former smoker | 32,119 | 396 | 1.21 (1.05, 1.38) |  |
| Current smoker | 24,821 | 351 | 2.05 (1.78, 2.37) |  |
| Smoking status unknown | 1163 | 21 | 1.44 (0.93, 2.23) |  |
| *Baseline smoking pack years* |  |  |  |  |
| 0-4 | 67,222 | 534 | 1 = reference | <2.22 x 10<sup>-16</sup> |
| 5-9 | 10,024 | 100 | 1.20 (0.97, 1.49) |  |
| 10-14 | 6868 | 46 | 0.98 (0.72, 1.32) |  |
| 15-24 | 10,445 | 156 | 1.62 (1.35, 1.93) |  |
| 25-49 | 10,106 | 195 | 1.75 (1.48, 2.06) |  |
| 50-99 | 2722 | 104 | 2.66 (2.15, 3.28) |  |
| 100+ | 135 | 3 | 1.79 (0.58, 5.58) |  |
| Unknown | 2846 | 61 | 1.90 (1.46, 2.48) |  |
| *Baseline alcohol status* |  |  |  |  |
| Never drink or less than once a week | 62,722 | 706 | 1 = reference |  |
| 1-2 drinks a week | 16,688 | 128 | 0.77 (0.63, 0.92) |  |
| 3-6 drinks a week | 15,788 | 137 | 0.79 (0.66, 0.95) |  |
| 7-10 drinks a week | 6637 | 94 | 1.13 (0.91, 1.40) | 0.0036 |
| 11-12 drinks a week | 1461 | 16 | 0.98 (0.60, 1.61) |  |
| 13-14 drinks a week | 1586 | 22 | 0.97 (0.63, 1.48) |  |
| More than 14 drinks a week | 2489 | 41 | 1.39 (1.01, 1.90) |  |
| Unknown | 2997 | 55 | 1.05 (0.80, 1.38) |  |
| *Baseline BMI* |  |  |  |  |
| <18.5 | 3420 | 35 | 1.56 (1.11, 2.20) |  |
| 18.5-24.9 | 67,552 | 587 | 1 = reference |  |
| 25.0-29.9 | 26,179 | 356 | 1.22 (1.07, 1.39) | 5.23 x 10<sup>-10</sup> |
| 30+ | 10,169 | 178 | 1.83 (1.54, 2.16) |  |
| Unknown | 3048 | 43 | 1.26 (0.93, 1.72) |  |
| *Diabetes<sup>a</sup>* |  |  |  |  |
| No | 85,497 | 761 | 1 = reference |  |
| Yes | 2856 | 79 | 2.64 (2.09, 3.33) | 4.75 x 10<sup>-12</sup> |
| Unknown | 22,015 | 359 | 1.03 (0.73, 1.46) |  |
| *Hypertension<sup>a</sup>* |  |  |  |  |
| No | 74,662 | 554 | 1 = reference |  |
| Yes | 13,691 | 286 | 1.78 (1.54, 2.05) | 4.86 x 10<sup>-13</sup> |
| Unknown | 22,015 | 359 | 1.15 (0.81, 1.63) |  |
| *Elevated serum cholesterol<sup>a</sup>* |  |  |  |  |
| No | 78,949 | 696 | 1 = reference |  |
| Yes | 9404 | 144 | 1.05 (0.88, 1.26) | 0.839 |
| Unknown | 22,015 | 359 | 0.98 (0.69, 1.39) |  |

<sup>a</sup>for persons responding to LQ2, with follow-up starting then

The radiation risks overall and by CeVD subtype are shown in Table 3 and Figure 1. There was little evidence of curvature in dose-response (*p*>0.1) for any endpoint. Cumulative thyroid dose was positively and linearly associated with deaths from overall CeVD (excess hazard ratio [EHR]/100 mGy = 0.07, 95% CI 0.01, 0.16, *p*=0.014). While all subtypes were positively associated with dose, only the subtype with the largest number of deaths, ischemic stroke (*n*=634 deaths) exhibited a significant dose-response (EHR/100 mGy = 0.10, 95% CI 0.02, 0.23, *p*=0.014). Additionally, ischemic stroke showed a larger dose-response trend than hemorrhagic stroke (*n*=369 deaths) (EHR/100 mGy = 0.04, 95% CI −0.04, 0.23 *p*=0.431) and other types of CeVD (*n*=196 deaths) (EHR/100 mGy = 0.02, 95% CI −0.10, 0.29, *p*=0.812). The differences in trend between the three subtypes were not significant (heterogeneity *p*=0.697). A similar dose-response trend was observed for total CeVD when modeling cumulative estimated badge dose rather than thyroid dose (EHR/100 mSv = 0.08, 95% CI 0.02, 0.16, *p*=0.005), also with little evidence of dose-response curvature (*p*=0.269) (Supplement A Table A2). A larger trend was observed using brain dose, with an EHR/100 mGy of 0.34 (95% CI −0.00, 0.81) (*p*=0.053), also with little evidence of curvature (*p*=0.507) (Supplement A Table A2). Supplement A Table A2 also reports the largely unadjusted analysis (adjusted only for sex), which yields a slightly higher EHR than for the fully-adjusted analysis reported there and in Table 3, with an EHR/100 mGy of 0.19 (95% CI 0.12, 0.28) (*p*<0.001); as previously there is little evidence of dose response curvature (*p*=0.458).

**Table 3.**
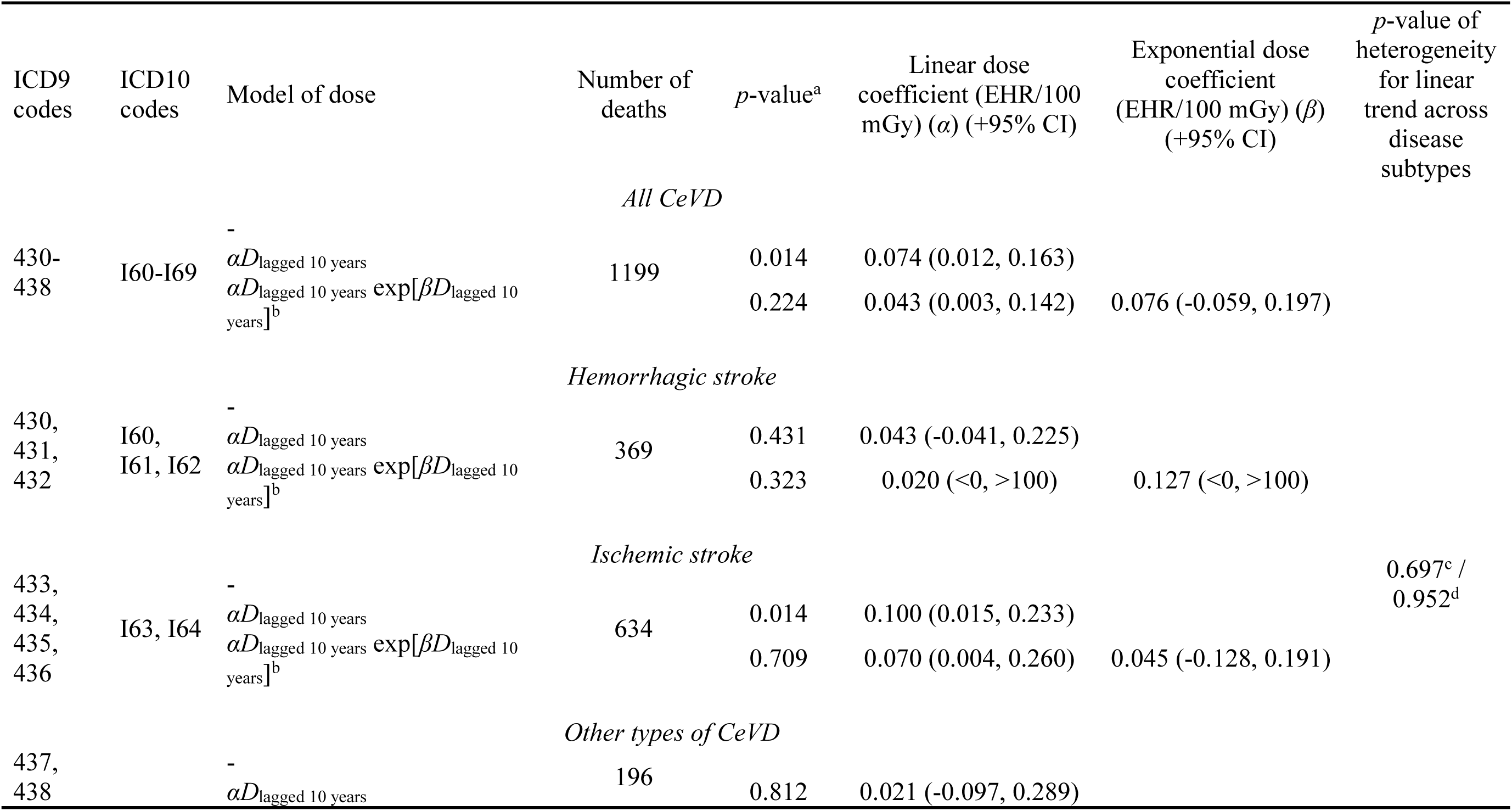
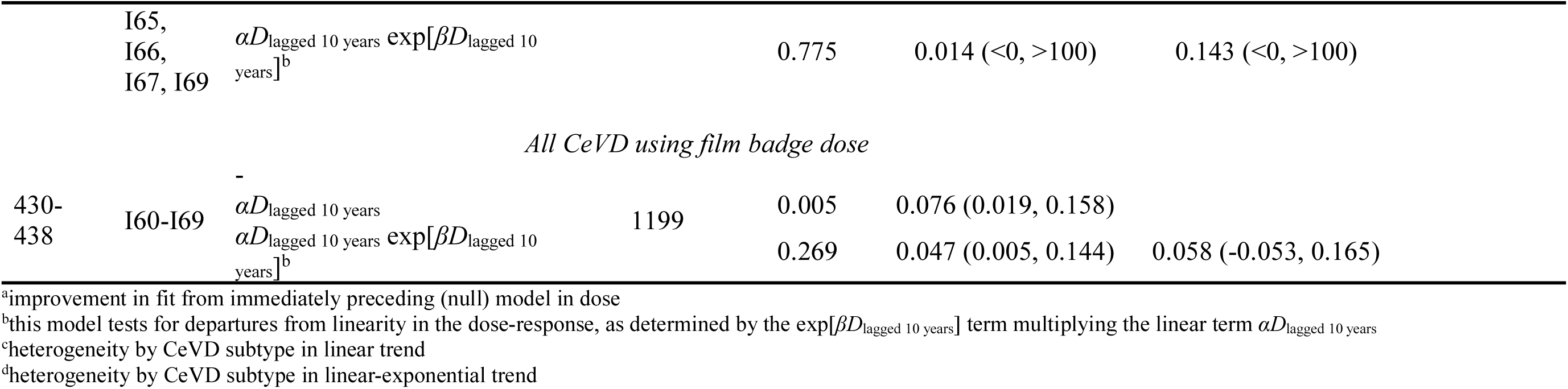
Results of fitting Cox proportional hazards model with cumulative dose lagged by 10 years to cerebrovascular disease (CeVD) mortality subtypes. Thyroid dose is used, with attained age at CeVD death ≤90. All models are adjusted for baseline smoking status, baseline pack years, birth year, baseline BMI, sex and baseline alcohol consumption by stratification

We explored using a slightly augmented set of adjusting variables, adding diabetes, elevated serum cholesterol, hypertension, using only cohort members who responded to LQ2 (Supplement A Table A3). These results indicate that these variables do not meaningfully alter the association between occupational radiation exposure and CeVD mortality. These variables were not retained in further analyses.

Table 4 shows the modifications in dose-response for various lifestyle variables. For BMI there was heterogeneity in response (*p*=0.037), with EHR largest for normal BMI (18.5-24.9 kg/m^2^) group. There was there no evidence of effect modification for any of the other variables (sex, smoking status, smoking pack-years, alcohol consumption) (*p*>0.15). Analysis of ischemic CeVD also suggested greater heterogeneity by BMI (*p*=0.010) and sex (*p*=0.054), with EHR largest for the normal BMI group (18.5-24.9 kg m^-2^) and for females.

**Table 4.** Modification of dose-response for all cerebrovascular disease (CeVD) and ischemic CeVD by lifestyle variables. Thyroid dose is used, with attained age at CeVD death ≤90. All models are adjusted for baseline smoking status, baseline pack years, birthyear, baseline BMI, sex, baseline alcohol consumption by stratification

| Variable used to modify | Dose coefficient<br>(EHR/100 mGy) ($\alpha$)<br>(+95% CI) | $p$-value of<br>heterogeneity | Dose coefficient<br>(EHR/100 mGy) ($\alpha$)<br>(+95% CI) | $p$-value of<br>heterogeneity |
| --- | --- | --- | --- | --- |
|  | *All CeVD* |  | *Ischemic CeVD* |  |
| Unmodified, total | 0.074 (0.012, 0.163) | NA | 0.100 (0.015, 0.233) | NA |
|  | *Sex* |  |  |  |
| Male | 0.018 (-0.040, 0.156) | 0.252 | -0.008 (-0.046, 0.137) | 0.054 |
| Female | 0.104 (0.022, 0.228) |  | 0.179 (0.049, 0.401) |  |
|  | *Baseline smoking status* |  |  |  |
| Unknown | 1.061 (-0.168, 18.59) | 0.399 | 1.171 (-0.186, 33.60) | 0.481 |
| Never smoked | 0.054 (-0.013, 0.170) |  | 0.105 (-0.004, 0.328) |  |
| Former smoker | 0.047 (-0.039, 0.211) |  | 0.029 (-0.050, 0.227) |  |
| Current smoker | 0.185 (0.002, 0.513) |  | 0.217 (-0.010, 0.755) |  |
|  | *Baseline smoking pack years* |  |  |  |
| Unknown and 0-4 | 0.064 (-0.003, 0.175) | 0.915 | 0.122 (0.012, 0.325) | 0.601 |
| 5-14 | 0.218 (-0.034, 0.902) |  | 0.428 (-0.026, 2.632) |  |
| 15-24 | 0.116 (-0.090, 0.662) |  | 0.117 (-0.095, 1.152) |  |
| 25-49 | 0.039 (-0.102, 0.347) |  | -0.032 (-0.123, 0.265) |  |
| 50+ | 0.064 (-0.047, 0.471) |  | 0.045 (-0.051, 0.502) |  |
|  | *Baseline alcohol consumption* |  |  |  |
| Unknown and <1 drink per week | 0.074 (0.005, 0.180) | 0.361 | 0.113 (0.016, 0.278) | 0.271 |
| 1-6 drinks per week | 0.164 (-0.036, 0.654) |  | -0.105 (-0.142, 0.306) |  |
| 7+ drinks per week | -0.075 (-0.120, 0.120) |  | -0.098 (-0.175, 0.146) |  |
|  | *Baseline BMI* |  |  |  |
| Unknown | -0.276 (-0.404, 0.213) | 0.037 | -0.286 (-0.406, 1.211) | 0.010 |
| <18.5 | 0.014 (-0.148, 1.463) |  | 0.109 (-0.147, 5.373) |  |
| 18.5-24.9 | 0.160 (0.049, 0.337) |  | 0.301 (0.103, 0.680) |  |
| 25-29.9 | 0.044 (-0.043, 0.221) |  | -0.024 (-0.064, 0.156) |  |
| 30+ | -0.096 (-0.144, 0.042) |  | -0.115 (-0.151, 0.030) |  |

There were no significant modifications to the dose trend for all CeVD for any of year of birth, time since exposure, age at exposure or attained age (*p*>0.2) (Table 5). Findings were similar for ischemic CeVD, although there was a significant (*p*=0.043) modification of risk by time since exposure: the dose trend appeared to increase up to 20-30 years after exposure, and then decrease.

**Table 5.** Best estimates of excess hazard ratio / Gy (+95% CI) by birth year and time windows of time since exposure, age at exposure and attained age in fit of linear relative risk (RR) model for cerebrovascular disease (CeVD) mortality and ischemic CeVD, with attained age at CeVD death ≤90. Analyses are adjusted for baseline smoking status, baseline pack years, birthyear, baseline BMI, sex and baseline alcohol consumption by stratification

|  | Excess hazard ratio /<br>100 mGy (+95% CI) | *p*-value<br>(heterogeneity) | Excess hazard ratio / 100 mGy<br>(+95% CI) | *p*-value (heterogeneity) |
| --- | --- | --- | --- | --- |
|  | *All CeVD* |  | *Ischemic CeVD* |  |
|  | Variation with birth year |  |  |  |
| <1910 | 0.315 (0.010, 4.744) | 0.733 | 0.220 (-0.013, 3.488) | 0.922 |
| 1910-1919 | 0.071 (-0.016, 0.240) |  | 0.075 (-0.024, 0.289) |  |
| 1920-1929 | 0.072 (-0.047, 0.277) |  | 0.095 (-0.056, 0.395) |  |
| 1930-1939 | 0.049 (-0.115, 0.303) |  | 0.082 (-0.151, 0.524) |  |
| 1940-1949 | -0.102 (-0.305, 0.229) |  | 0.089 (-0.282, 0.907) |  |
| 1950+ | 0.005 (-0.540, 1.385) |  | 1.270 (-0.436, 45.69) |  |
|  | Variation with time since exposure |  |  |  |
| 10-20 years before | -0.736 (-1.334, 0.157) | 0.197 | -1.092 (-1.579, -0.077) | 0.043 |
| 20-30 years before | 0.352 (-0.299, 1.156) |  | 1.135 (0.035, 2.585) |  |
| 30+ years before | 0.079 (0.015, 0.171) |  | 0.090 (0.009, 0.220) |  |
|  | Variation with age at exposure |  |  |  |
| <25 years | 0.085 (0.018, 0.182) | 0.588 | 0.101 (0.014, 0.241) | 0.420 |
| 25-34 years | 0.281 (-0.418, 1.284) |  | 0.937 (-0.338, 3.033) |  |
| 35-49 years | -0.147 (-0.515, 0.334) |  | -0.331 (-0.737, 0.325) |  |
| 50+ years | -0.127 (-0.566, 0.489) |  | 0.265 (-0.495, 1.300) |  |
|  | Variation with attained age |  |  |  |
| <70 years | 0.121 (-0.081, 0.511) | 0.720 | 0.076 (-0.116, 0.704) | 0.894 |
| 70+ years | 0.070 (0.008, 0.162) |  | 0.102 (0.014, 0.245) |  |

Supplement A Table A4 shows the significance of dose-response trend by range of thyroid dose used. Using 1-sided tests of significance the trend over the cumulative thyroid dose range 0-1000 mGy was statistically significant (EHR/100 mGy=0.07, 90% CI 0.01, 0.14, 1-sided *p*=0.029), but significance was lost (*p*>0.05) for any lower range of dose. Varying the lag period between 5 and 20 years yielded little change in EHR (Supplement A Table A5).

## Discussion

In this large cohort of U.S. radiologic technologists, we observed a significant linear trend of CeVD mortality with cumulative thyroid absorbed dose as well as brain and film badge dose. The CeVD subtype with the largest number of deaths, ischemic stroke, was the only subtype to exhibit a significant dose-response (*p*=0.014), and exhibited the steepest trend with dose; however, there were no significant (*p*>0.5) differences in trend between the three CeVD subtypes. Findings suggested an association with dose range of 0-1000 mGy. There were indications of effect modifications by sex and BMI with ischemic CeVD, with largest risk estimates in female workers and workers with a normal BMI (18.5-24.9 kg m^-2^), but no significant modifications of trends with year of birth, time since exposure, age at exposure, or attained age for all CeVD.

To our knowledge, this is the first study of radiation workers that examined CeVD mortality risk by subtype and identified a dose-response relationship with exposure to ionizing radiation. Only three previous studies assessed CeVD subtype. In the cohort of Mayak plutonium production workers,^23^ the ERRs were not significant for any subtype (Supplement A Table A6). It was also examined in the Semipalatinsk study of persons exposed to atmospheric nuclear tests in the former USSR,^24^ which found a strong and significant ERR for ischemic stroke, and a non-significant one for hemorrhagic stroke, although the central estimates for the two endpoints were comparable. The French study of childhood cancer survivors who had been treated with radiotherapy^25^ found a notable ERR for ischemic stroke which was stronger than for overall stroke. Both the Japanese atomic bomb survivors cohort^26^ and the large three-country International Nuclear Worker Study (INWORKS)^5^ observed significant radiation-related dose-response trends for overall CeVD, but neither of these studies reported risks by CeVD subtype. Our finding for ischemic stroke mortality is in line with previously-reported stronger associations between ionizing radiation and ischemic heart disease compared to other types of heart disease.^3^ While pathophysiological mechanisms for vascular effects of ionizing radiation at low doses are speculative, potential mechanisms may include endothelial damage and dysfunction, changes to the arterial intima, vascular inflammation and resulting atherosclerotic changes.^3,27,28^ While there is some overlap with mechanisms resulting in hemorrhagic and other CeVD mortality, these vascular changes are much more pronounced in ischemic disease.

Our finding of few indications of modification of dose-response by demographic, lifestyle and other morbidity risk factors is consistent with the small number of studies of CeVD examined in a recent systematic review.^3,29^ The weak non-significant modifications in dose-response by time since exposure, attained age and age at exposure for CeVD overall were consistent in direction with those identified in a recent review.^30^

Other studies of CeVD mortality and morbidity, which we summarise in Supplement A Table A6 show modest trends. A similar magnitude of the EHRs per unit radiation dose was derived using estimated film badge or thyroid-absorbed dose. This is not surprising because the thyroid gland is generally not protected by lead aprons and most medical radiation workers do not use thyroid shields. Most other occupational studies of radiation workers evaluated film badge dose in relation to CeVD risk, and the trends in ERRs they derive are similar to our analysis EHRs using film badge dose.

Strengths of the present study include the large population with low-to-moderate cumulative, protracted occupational radiation exposure, large size and prospective cohort design, and opportunity to evaluate risks separately for female and male radiation workers. We utilised a state-of-the-art occupational dosimetry with estimated absorbed doses specific to the thyroid, brain and badge, which is relevant because brain doses are much lower than thyroid doses (Table 1). Although there were multiple sources of uncertainty in the reconstruction of estimated cumulative occupational dose,^7^ earlier dose estimates were found to be associated with chromosome translocation rate in a study using fluorescence *in situ* hybridisation (FISH).^31^ The comprehensive demographic, lifestyle, and medical history data are major strengths of our study, as the availability of these data provided a rare opportunity among occupational radiation cohort studies to evaluate confounding, and effect modification by work history factors and major risk factors for CeVD. The fact that there are so few indications of confounding or effect modification by these variables provides support that the results presented may be causal. Another novelty and strength of the present study is the analysis of time-dependent doses accumulated in windows of time since exposure, age at exposure, and attained age; to the best of our knowledge only the INWORKS pooled nuclear worker cohort,^5^ the LSS ^26,32^ and a study of persons treated with X-ray for peptic ulcer^13^ have assessed these variables for CeVD.

However, our study had several limitations. The medical and lifestyle variables evaluated as potential confounders and modifiers were ascertained solely by questionnaire and were not validated. However, the population of radiologic technologists reported here is medically literate, so that self-reporting of health and medical risk factors should be reasonably reliable. The effect of most of these variables on CeVD risk (Table 2) is similar to those reported in other cohorts^2^, suggesting reasonable ascertainment. The lifestyle and medical data were collected at the first or second questionnaire, but the radiation exposures for many, and in particular all the larger radiation doses, will have occurred many years previously, so the lifestyle/medical information cannot be regarded as truly baseline; thus, the ‘baseline’ characteristics reported in the first or second questionnaire may have occurred after the protracted radiation exposure, or may have changed during follow-up. Follow-up occurred via mortality rather than incidence. However, the relatively high positive predictive value (PPV) of stroke diagnosis via death certification in the United States, of ∼80% or more,^10^ suggests that in our cohort the vast majority of stroke deaths identified as underlying cause via death certificate will truly be due to stroke. Nonetheless, the reliance on cause-of-death information meant that individuals who were diagnosed but did not die due to stroke during the follow-up period were not included in the outcome definition. Our results may capture radiation-associated risks for only the more severe forms of stroke, and are less applicable to individuals more likely to survive following a stroke event. Future analyses of occupational radiation exposure with stroke incidence may reveal some differences in the magnitude or shape of the dose-response relationship.

## Conclusions

In summary, analysis of CeVD mortality in the USRT cohort suggests a trend with dose, which was more pronounced for ischemic CeVD than other CeVD subtypes. There was no suggestion of non-linearity in dose-response. This cohort is unique among occupational groups with regard to the lifestyle and medical risk factor data available. Generally year of birth, time since exposure, age at exposure and attained age did not modify the CeVD dose-response, nor did sex or lifestyle/medical risk factors, including smoking, alcohol consumption, BMI, diabetes, elevated serum cholesterol, or hypertension. However, for ischemic CeVD, there were indications of modification by time since exposure, sex, and BMI. These novel findings, particularly in relation to modification of the strong linear trends with dose for ischemic CeVD, require replication and further investigation to understand biological mechanisms. These results reinforce the need for radiation protection practices aimed at keeping exposures as low as reasonably achievable.

## Data Availability

Data are available upon reasonable request. These data are available from the National Cancer Institute, Division of Cancer Epidemiology and Genetics, and upon submission and approval of a research proposal and completion of a Data Transfer Agreement.

## Acknowledgments

The USRT study is funded by the Intramural Research Program of the National Institutes of Health, the National Cancer Institute, Division of Cancer Epidemiology and Genetics. The contributions of the NIH authors were made as part of their official duties as NIH federal employees, are in compliance with agency policy requirements, and are considered Works of the United States Government. However, the findings and conclusions presented in this paper are those of the authors and do not necessarily reflect the views of the NIH or the U.S. Department of Health and Human Services. The authors thank the radiologic technologists who participated in the study. The authors would also like to thank Allison Iwan of the University of Minnesota and the American Registry of Radiologic Technologists for their continued support.

## Ethical approval

This study has been approved annually by the University of Minnesota Institutional Review Board.

## Supplement A

**Table A1.** Multivariable analysis of cerebrovascular disease (CeVD) in relation to variables deemed significant from the univariable analysis of Table 2. Analysis using a Cox proportional hazards model with age as timescale. Variables are added in order of significance from the univariable analysis of Table 2.

| Variables used to model hazard ratio | *p*-value for improvement in fit of model on previous line |
| --- | --- |
| Baseline smoking status | $<2.22 \times 10^{-16}$ <sup>a</sup> |
| Baseline smoking status + baseline pack years | $2.05 \times 10^{-8}$ |
| Baseline smoking status + baseline pack years + birthyear | $1.14 \times 10^{-9}$ |
| Baseline smoking status + baseline pack years + birthyear + baseline BMI | $6.77 \times 10^{-11}$ |
| Baseline smoking status + baseline pack years + birthyear + baseline BMI + sex | $3.34 \times 10^{-3}$ |
| Baseline smoking status + baseline pack years + birthyear + baseline BMI + sex + baseline alcohol | 0.0428 |
| Baseline smoking status + baseline pack years + birthyear + baseline BMI + sex + baseline alcohol + hypertension | $2.91 \times 10^{-9}$ <sup>b, c</sup> |
| Baseline smoking status + baseline pack years + birthyear + baseline BMI + sex + baseline alcohol + hypertension + diabetes status | $7.66 \times 10^{-7}$ <sup>c</sup> |

<sup>a</sup>*p*-value of improvement in fit over null model
<sup>b</sup>*p*-value of improvement in fit over model with baseline smoking status + baseline pack years + birthyear + baseline BMI + sex + baseline alcohol fitted to data for persons responding to LQ2, with follow-up starting then
<sup>c</sup>fitted to data for persons responding to LQ2, with follow-up starting then

**Table A2.**
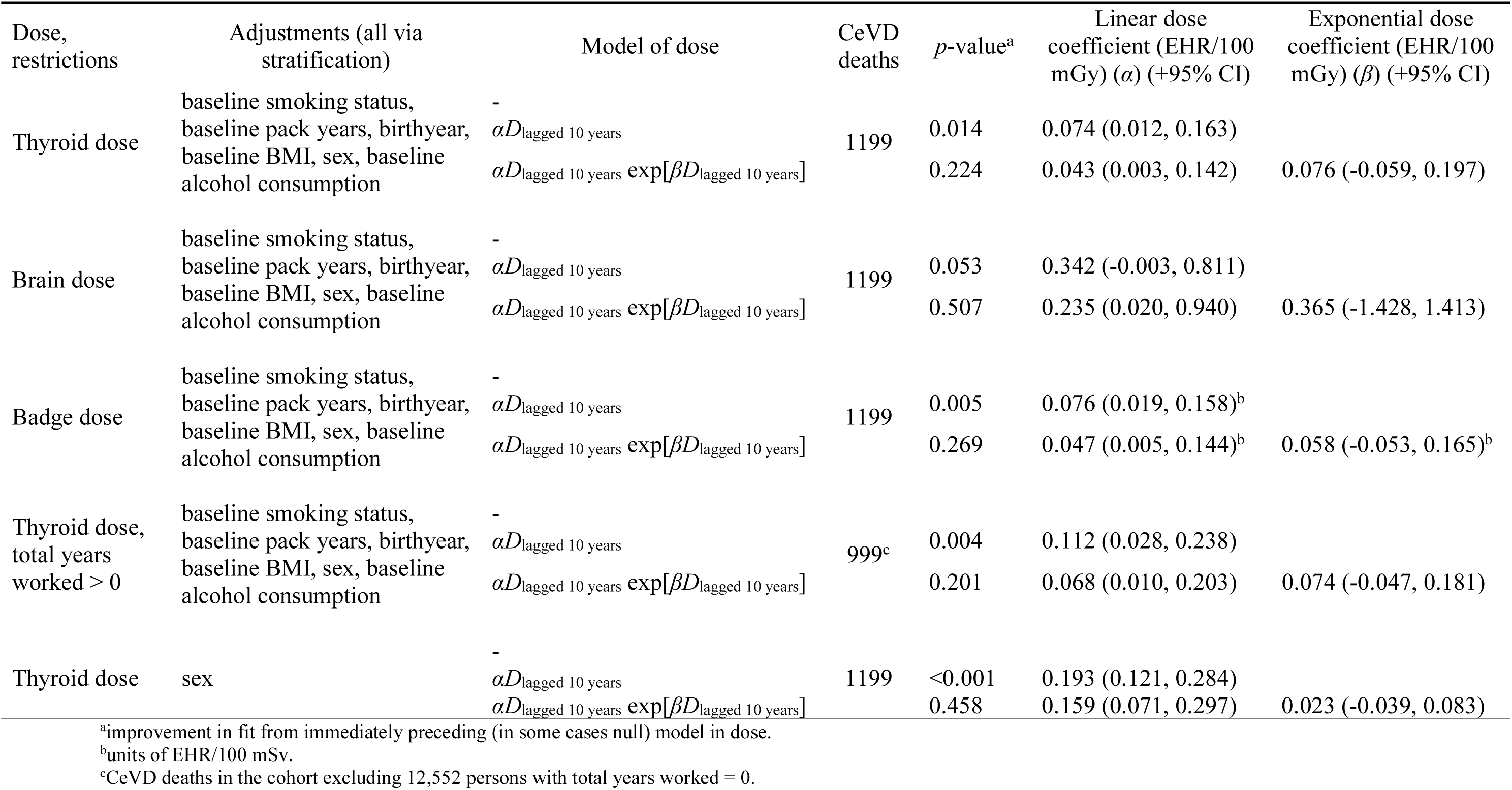
Results of fitting Cox proportional hazards model with cumulative dose lagged by 10 years to cerebrovascular disease (CeVD) mortality. Thyroid dose, badge dose and brain dose are used. Most models are adjusted for baseline smoking status, baseline pack years, birthyear, baseline BMI, sex, baseline alcohol consumption by stratification, one only for sex.

**Table A3.** Results of fitting Cox proportional hazards model with cumulative thyroid dose lagged by 10 years to cerebrovascular disease (CeVD) mortality, with attained age at CeVD death ≤90, and additional adjustment for medical history. All analyses fitted to data for persons responding to LQ2, with follow-up starting then, and are adjusted for baseline smoking status, baseline pack years, birthyear, baseline BMI, sex and baseline alcohol consumption by stratification

| CeVD deaths | Baseline stratified model |  |  | Additional adjustment via stratification |  |  |
| --- | --- | --- | --- | --- | --- | --- |
|  | Dose-response model | *p*-value<sup>a</sup> | Excess hazard ratio / 100 mGy (+95% CI) | Additional stratification variable | *p*-value<sup>a</sup> | Excess hazard ratio / 100 mGy (+95% CI) |
| 873 | - |  |  |  |  |  |
| | $\alpha D_{\text{lagged 10 years}}$ | 0.175 | 0.062 (-0.023, 0.186) | Diabetes | 0.487 | 0.033 (-0.049, 0.154) |
|  | - |  |  |  |  |  |
| | $\alpha D_{\text{lagged 10 years}}$ | 0.175 | 0.062 (-0.023, 0.186) | Elevated serum cholesterol | 0.154 | 0.070 (-0.021, 0.204) |
|  | - |  |  |  |  |  |
| | $\alpha D_{\text{lagged 10 years}}$ | 0.175 | 0.062 (-0.023, 0.186) | Hypertension | 0.468 | 0.034 (-0.046, 0.153) |

<sup>a</sup>improvement in fit from immediately preceding (null) model in dose

**Table A4.** Dose-response trend by range of dose used. 10-year lagged thyroid dose is used, with attained age at cerebrovascular disease (CeVD) death ≤90. All models are adjusted for baseline smoking status, baseline pack years, birthyear, baseline BMI, sex, baseline alcohol consumption by stratification

| Dose limit mGy | CeVD Deaths | Dose coefficient (EHR/100 mGy) ($\alpha$) (90% CI) | 1-sided $p$-value for trend | Dose coefficient (EHR/100 mGy) ($\alpha$) (95% CI) | 2-sided $p$-value for trend |
| --- | --- | --- | --- | --- | --- |
| Unlimited | 1199 | 0.074 (0.020, 0.147) | 0.007 | 0.074 (0.012, 0.163) | 0.014 |
| <1500 | 1192 | 0.062 (0.008, 0.135) | 0.026 | 0.062 (-0.001, 0.151) | 0.053 |
| <1250 | 1186 | 0.056 (0.001, 0.128) | 0.046 | 0.056 (-0.008, 0.144) | 0.092 |
| <1000 | 1181 | 0.065 (0.007, 0.141) | 0.029 | 0.065 (-0.002, 0.158) | 0.058 |
| <750 | 1169 | 0.056 (-0.005, 0.135) | 0.069 | 0.056 (-0.015, 0.152) | 0.138 |
| <500 | 1131 | 0.008 (-0.056, 0.090) | 0.425 | 0.008 (-0.066, 0.108) | 0.850 |
| <400 | 1102 | -0.028 (-0.095, 0.057) | 0.277 | -0.028 (-0.106, 0.076) | 0.553 |
| <300 | 1058 | 0.000 (-0.087, 0.109) | 0.498 | 0.000 (-0.101, 0.133) | 0.996 |
| <200 | 951 | -0.058 (-0.169, 0.082) | 0.233 | -0.058 (-0.187, 0.113) | 0.467 |
| <100 | 679 | -0.241 (-0.438, 0.016) | 0.060 | -0.241 (-0.471, 0.074) | 0.120 |

**Table A5.** Results of fitting Cox proportional hazards model with cumulative thyroid dose lagged by varying numbers of years to cerebrovascular disease (CeVD) mortality, with attained age at CeVD death ≤90. Analyses are adjusted for baseline smoking status, baseline pack years, birthyear, baseline BMI, sex and baseline alcohol consumption by stratification

| Lag period, years | -2 x log [partial likelihood]<sup>a</sup> | *p*-value<sup>b</sup> | Linear dose coefficient (EHR/100 mGy) ($\alpha$) (+95% CI) |
| --- | --- | --- | --- |
| 5 | 8796.58 | 0.017 | 0.071 (0.010, 0.158) |
| 10 | 8796.25 | 0.014 | 0.074 (0.012, 0.163) |
| 15 | 8795.89 | 0.012 | 0.078 (0.014, 0.168) |
| 20 | 8795.72 | 0.011 | 0.080 (0.015, 0.172) |

<sup>a</sup>-2 x log[partial likelihood] a standard measure of goodness of fit for the Cox model; the smaller this number is, the better the fit
<sup>b</sup>improvement in fit from null model in dose

**Table A6.**
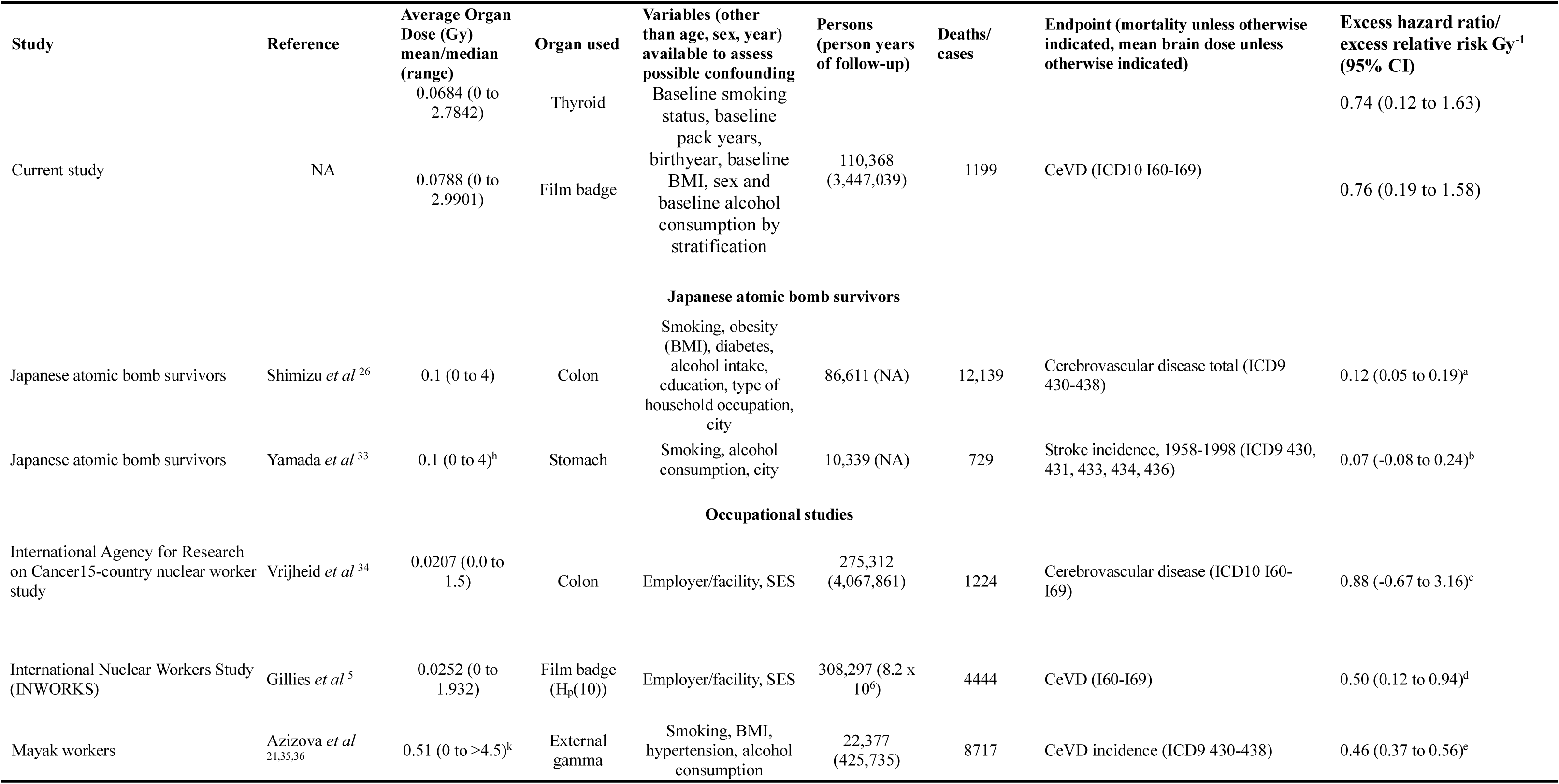
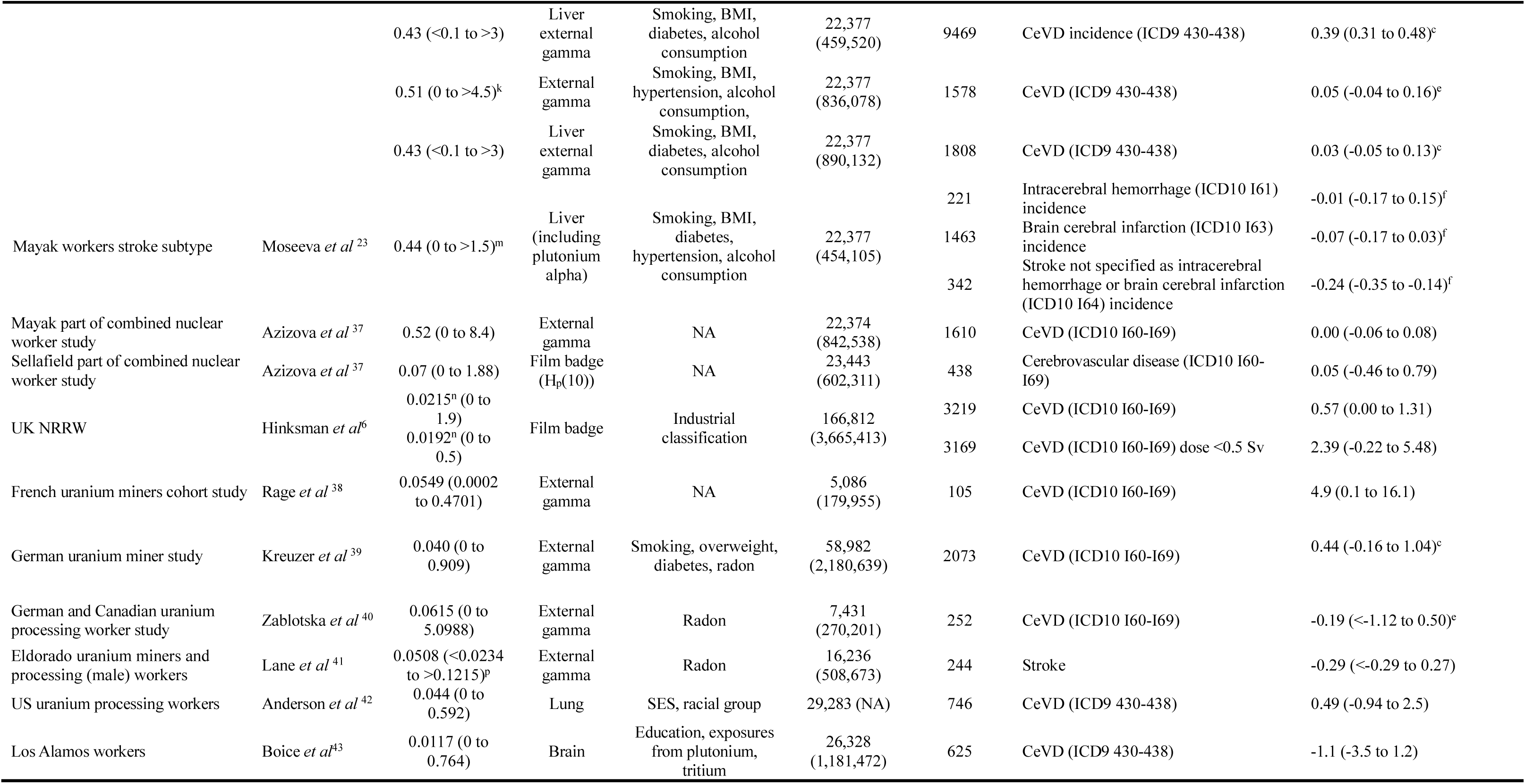
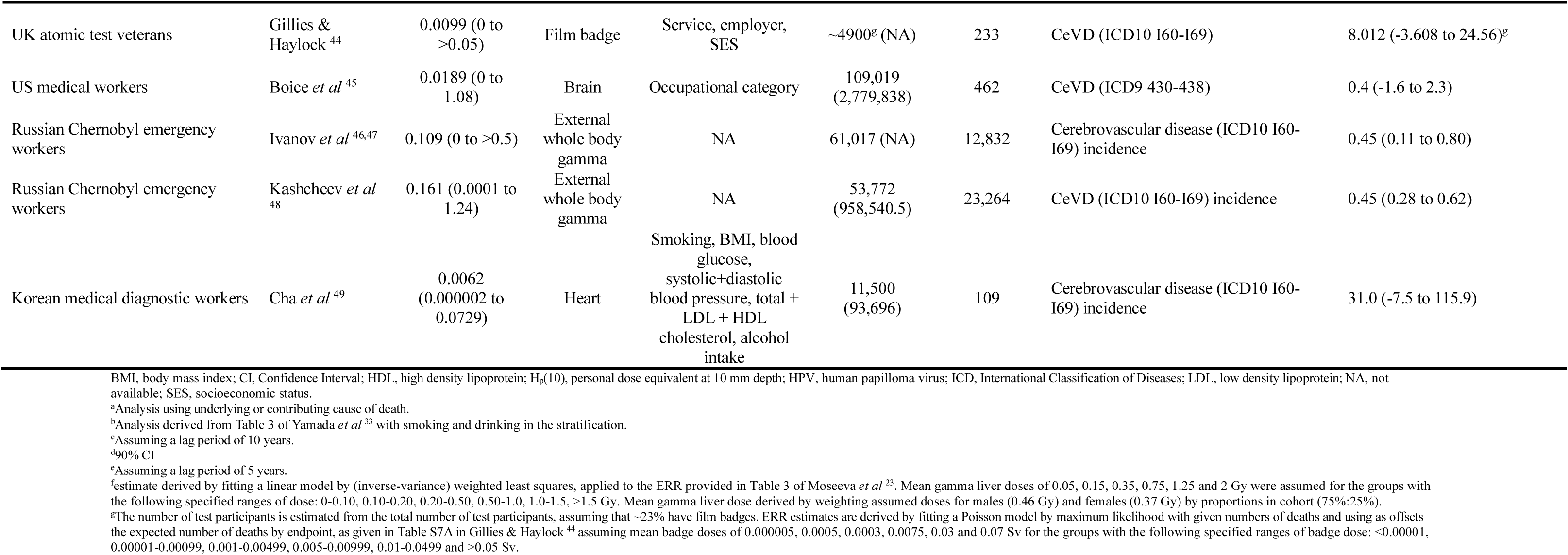
Estimated excess relative risk of cerebrovascular disease (CeVD) in selected occupational studies and Japanese atomic bomb survivors. All data are in relation to underlying cause of death, unless otherwise indicated. Wherever possible 5 year lag is used. Only studies with >100 cases or deaths are shown.

## Supplement B. Supplementary Statistical Methods and Results

### Statistical methods

Mortality risks in the cohort were assessed using Cox proportional hazards models,^20^ with age as timescale, in which the relative risk (RR) for individual *i* at age *a* was given by:

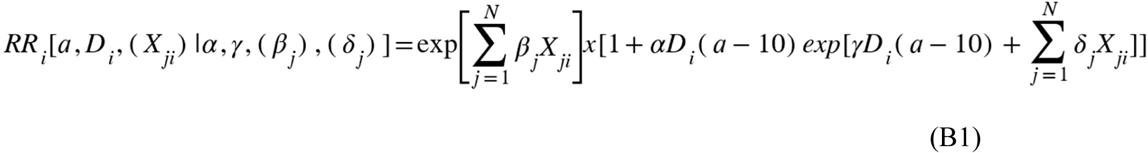

where *D_i_*(*a*–10) is the time-varying cumulative dose in mGy, lagged by 10 years to allow for disease latency,^32^ (*X_ji_*) are the cardiovascular risk factors (other than occupational radiation exposure), *α* is the excess relative risk coefficient (EHR) per unit dose (mGy), *γ* is the exponential coefficient in dose (to test for curvature in the dose-response), and (*β_j_*) and (*δ_j_*) are coefficients adjusting for lifestyle risk factors. We report the –2 x log[partial likelihood] statistic in various tables, a standard measure of goodness of fit for the Cox model; since we maximize the partial likelihood in model fitting, the smaller this number is, the better the fit. We evaluated several dose metrics including thyroid dose (a surrogate for dose to the carotid artery), whole brain dose, and film badge dose. We also explored varying the dose lag period between 5 and 20 years. Because of the possible unreliability of cause of death ascertainment at advanced ages,^10^ we performed sensitivity analyses in which subsets of the data with maximum age of CeVD deaths were progressively reduced, to 100, 90 and 85 years. For most analyses we used a maximum age of CeVD of 90 years. Because of suspicions that the group of technologists who were certified but did not actually work as radiologic technologists may be at increased risk of CeVD and other diseases, as would be expected from the healthy worker selection effect^50^ among other things, further sensitivity analysis was conducted restricting to those with more than 0 self-reported years worked as a radiologic technologist, resulting in the removal of 12,552 technologists from the analysis dataset. We selected the set of covariates used via a multivariable analysis (as yielding significant modification of CeVD risk), specifically baseline smoking status, baseline pack years, birthyear, baseline body mass index (BMI), sex and baseline alcohol consumption (for details see Table 2, Supplement A Table A1); for most analysis we used only these variables to adjust the models, by stratification. We assessed modifications to the dose trend by time since exposure, age at exposure and attained age. We explored adding adjustment for some variables, in particular diabetes, elevated serum cholesterol and hypertension, which may be on the pathway between ionizing radiation exposure and CeVD mortality, and may also (by virtue of their appearance on second, but not first, mailed questionnaires) be markers of survival to the later periods of follow-up. Analyses using these variables (diabetes, elevated serum cholesterol, hypertension) were restricted to those answering the second questionnaire (LQ2) (when the relevant questions were asked) and starting follow-up at the date of completion of the questionnaire.

### Results

Table 2 shows the univariate associations for various baseline variables with CeVD mortality, most of which were highly significant (*p*<0.001), except for race, ethnicity and elevated serum cholesterol (restricted to LQ2 follow-up). Supplement A Table A1 shows the results of a stepwise analysis, adding the variables in order of the *p*-values from the univariate analysis of Table 2. Most variables from the primary set (baseline smoking status, baseline pack years, birthyear, baseline BMI, sex) remained highly significant in this stepwise analysis (*p*<0.005), but the significance of baseline alcohol was markedly reduced (*p*=0.0428).

Supplement A Table A2 shows the results of fitting a linear Cox model using 10-year lagged cumulative thyroid dose, brain dose and film badge dose. A linear association was observed for thyroid dose, with an excess hazard ratio (EHR) /100 mGy of 0.07 (95% CI 0.01, 0.16) (*p*=0.014), but only of borderline significance for brain dose, with an EHR/100 mGy of 0.34 (95% CI −0.00, 0.81) (*p*=0.053) and film badge dose, with an EHR/100 mSv of 0.08 (95% CI 0.02, 0.16) (*p*=0.005). The EHR did not markedly change after reducing the maximum attained age at CeVD death to 100, 90 or 85 years (results not shown). As with thyroid dose, there was little evidence of dose-response curvature (*p*>0.2) using film badge or brain dose. Restricting the analysis to those with total years worked greater than 0, the number of CeVD deaths decreased from 1199 to 999, and the EHR/100 mGy thyroid dose increased slightly, to 0.11 (95% CI 0.03, 0.24) (*p*=0.004).

We further explored varying the lag period between 5 and 20 years. The EHR/Gy was slightly larger, and more statistically significant, with use of longer lags (Supplement A Table A3). Analysis was conducted using R ^22^ and Fortran 95-2003 code written by the first author.

**Figure 1.**
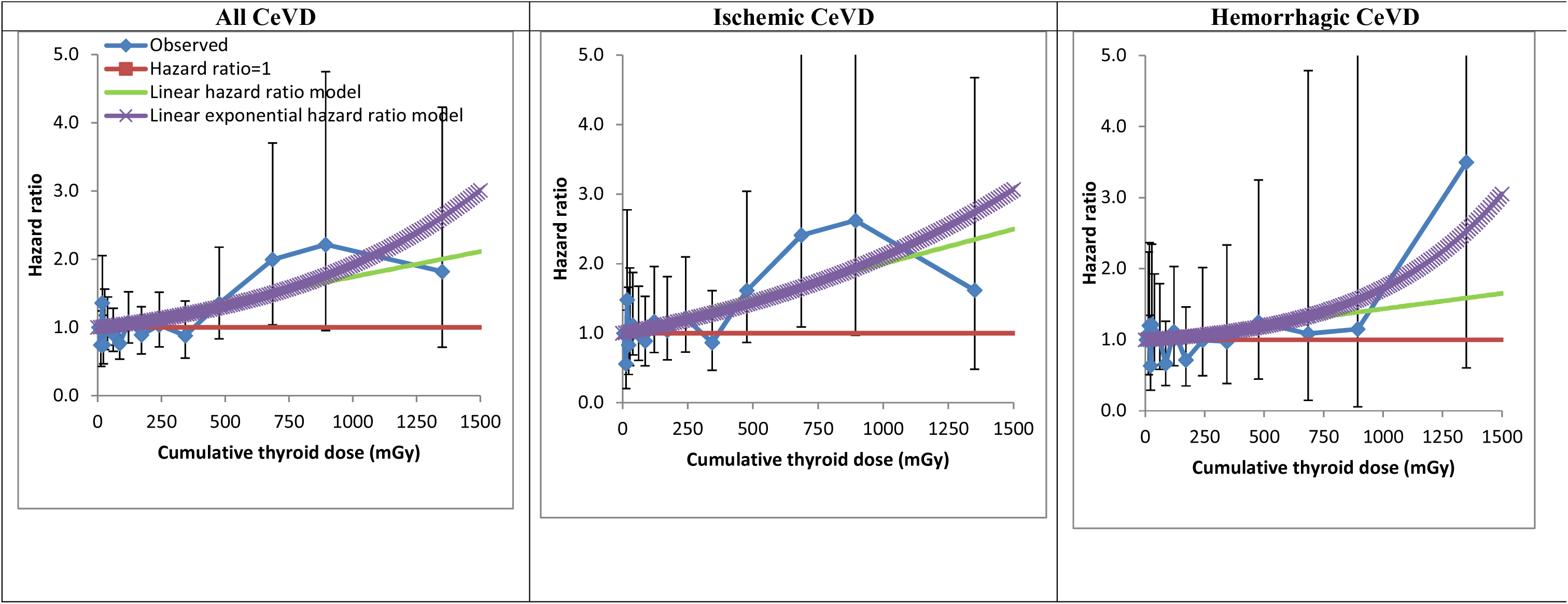
Dose-response (+95% CI) for all cerebrovascular disease (CeVD), ischemic CeVD and hemorrhagic CeVD in relation to 10-year-lagged cumulative thyroid dose. Analysis is of persons with CeVD deaths restricted to ≤90, adjusted for baseline smoking status, baseline pack years, birthyear, baseline BMI, sex and baseline alcohol consumption via stratification.

